# Reliability and equivalence of the 8-item version of the Patient Health Questionnaire (PHQ-8) to screen for depressive symptoms in adults with chronic pain: A representative U.S. population study

**DOI:** 10.1101/2025.08.08.25333327

**Authors:** Jennifer S. De La Rosa, Greg T. Chism, Katherine E. Herder, Chung Jung Mun, Rachel V. Aaron

## Abstract

**Background:** The 8-item Patient Health Questionnaire (PHQ-8) is the most widely implemented clinical depression screener in the world and is increasingly influential in population surveillance of unmet mental health needs. However, in the context of chronic pain (CP), depression screening could be complicated by overlapping symptoms. Inaccurate attribution of CP symptoms to depression could lead to overestimation of depressive symptom severity or even artifactual inflation of depression prevalence estimates. Results of small, non-representative studies are heterogenous; suitability of the PHQ-8 to screen for depression in those with CP remains unclear. We aimed to determine the reliability and equivalence of PHQ-8 in adults with and without CP.

**Methods:** This cross-sectional observational population study used representative data from the 2019 National Health Interview Survey. Descriptive statistics and visualizations were generated; reliability, cross-group equivalence and measurement invariance were assessed.

**Findings:** The final sample contained 30,983 U.S. adults. Prevalence of clinically significant depressive symptoms was 3.5% in those without CP, 20.1% in those with CP, and 34.8% in those with high-impact chronic pain (HICP). Reliability, measurement invariance and cross-group equivalence were observed at the configural, metric, and scalar levels. No evidence consistent with overestimation of depression prevalence or severity in the contexts of CP or HICP was observed.

**Interpretation:** The PHQ-8 is reliable and measures depressive symptoms equivalently in the context of CP, supporting the continued use of PHQ-8 to screen for depression in clinical and research settings and in population surveillance of adults with and without CP. CP was associated with five times higher prevalence of clinically significant depressive symptoms; findings suggest this reflects true cross-group differences in prevalence and severity of depressive symptoms and is unlikely to result from measurement issues.

**Funding:** This work was funded in part by the Comprehensive Center for Pain & Addiction, University of Arizona, and by NIH K23HD104934.

**RESEARCH IN CONTEXT:** 

**Evidence before this study:** PubMed and Google Scholar were searched without time constraints using Key Terms: “depress∗” and (“chronic pain” OR “fibromyalgia” OR “migraine” OR “headache” OR musculoskeletal” OR “arthritis”) and “assess∗” OR (“measur∗” OR “evaluat∗ OR “screen*”). Additional studies were identified from the reference lists of the resulting papers. Though many studies have examined assessment of depression in the context of chronic pain, nearly all appear to be limited by non-representative samples, failure to incorporate non-focal comparison groups, or both. As a result, the reliability and equivalence of standard depression screening tools to measure depressive symptoms in the clinical context of chronic pain has remained persistently unclear.

**Added value of this study:** The PHQ-8 is the most widely implemented depression screening tool in the world: it is frequently used in clinical practice, scientific research, and is becoming increasingly influential in the global surveillance of unmet mental health needs. Systematic bias of depression screening in the context of chronic pain is an important concern. If PHQ-8 functions differently in those with chronic pain, clinical decisions, scientific studies, and even epidemiological estimates could be substantially biased. To our knowledge, this cross-sectional survey study of 30,983 U.S. adults is the first to use representative population data comparing those with and without chronic pain to determine the reliability and cross-group equivalence of the PHQ-8: addressing a persistent knowledge gap with important implications for clinical practice, scientific research, and global public health. The PHQ-8 was demonstrated to be reliable in adults with chronic pain. Cross-group equivalence and measurement invariance were observed between those with and without chronic pain at the configural, metric, and scalar levels. These results provide compelling evidence that PHQ-8 is reliable and functions equivalently to screen for depression in adults with and without chronic pain.

**Implications of all the available evidence:** Findings should enhance confidence among clinicians and researchers that PHQ-8 is appropriate to screen for depressive symptoms in the clinical context of chronic pain and that the interpretation of screening results is comparable and straightforward. The available evidence does not support the hypothesis that measurement artifacts contribute to elevated rates of positive PHQ-8 screenings in those with chronic pain. On the contrary, the findings underscore the existence of the profound mental health disparities experienced by U.S. adults with chronic pain from end-to-end across the patient journey, including: higher prevalence of clinically significant depressive symptoms, a lower likelihood that such symptoms will be identified, and a lower likelihood of receiving needed mental health support.

## 1. INTRODUCTION

Depression and chronic pain (CP) commonly co-occur in general and patient populations. Among adults who screen positive for depression, the majority (an estimated 56%) have co-occurring CP,^1^ while approximately 40% of adults with CP have co-occurring depression.^2^ About half of these—an estimated 20% of adults with CP—have clinically significant depressive symptoms that have yet to be effectively addressed.^1^ The co-occurrence of CP and depressive symptoms is associated with substantially higher prevalence of functional limitations in work, daily activities, and social participation, compared to those with CP alone or with depressive symptoms alone.^1^ Addressing the mental health needs of those with CP is essential to optimal functioning and quality of life,^3^ yet those with CP have 40% lower odds of receiving needed mental health treatment.^4^ Addressing the unmet mental health needs of people with CP is recognized as a key priority for global population health, and as a fundamental human right.^5,6^ It is worth noting explicitly that our conceptualization of unmet mental health needs is in no way exclusive of emotional distress generated by (or intensified by) the life-altering experience of chronic pain itself. Therefore, to address the unmet mental health needs of those with chronic pain and clinically significant depressive symptoms, expanded access to pain care *and* mental health care is urgently needed.

Depression screening tools have been implemented into general health settings worldwide; today, most cases of major depression are identified and addressed in primary or urgent care settings.^7^ CP frequently co-occurs with depression, and CP may be especially prevalent in settings where depression screeners are implemented into routine clinical practice. However, depression screening could be confounded by the fact that several overlapping somatic symptoms commonly occur with CP, such as sleep disturbance, fatigue, appetite changes, and psychomotor slowing, that are also diagnostic criteria for depression. Furthermore, concerns about criterion contamination need not be restricted to somatic symptoms: the International Association for the Study of Pain’s definition of pain explicitly presumes the accompaniment of negative affective states.^8^ Criterion contamination is an important concern, in that if CP symptoms were inaccurately attributed to depression, this could result in overestimation of depressive symptom severity or even artifactual inflation of depression prevalence estimates among those with CP.^9,10^

Several prior studies have explored the validity of self-report depression screeners in the context of CP.^11^ However, the majority have been limited by the use of small, non-representative samples, lack of non-focal comparison groups (i.e., persons without CP and persons without depression), or both. Findings are heterogenous, with some studies demonstrating acceptable validity of depression screening tools in those with CP^12,13^ and others finding support for psychometric concerns.^14,15^

The Patient Health Questionnaire-8 (PHQ-8) is one of the most frequently implemented depression screeners worldwide, further, epidemiological estimates based on the PHQ-8 are used to support development of mental health policy, including the allocation of resources to health services and research.^16^ Therefore, the possibility that PHQ-8 could be systematically biased in the context of CP warrants rigorous investigation. While the PHQ-8 has demonstrated excellent validity and reliability across diverse geographies, settings, and contexts,^17^ its equivalence in the context of CP remains unclear. If PHQ-8 functions differently in CP, clinical decisions, scientific research, and prevalence estimates could be biased. This study used representative U.S. population data to evaluate the reliability and cross-group equivalence of PHQ-8 to screen for depressive symptoms in the context of CP, addressing a persistent knowledge gap with important implications for clinical practice, research, and global public health.

## 2. METHODS

### 2.1 Data

The 2019 National Health Interview Survey (NHIS) data (n=31,997) is sampled and weighted to represent 244.6 million adults in the U.S. population.^18^ NHIS data are routinely used to estimate prevalence, incidence, trends, and disparities related to depressive symptoms and CP.^19–21^

### 2.2 Measures

**Chronic Pain (CP)** was measured using the item: “*In the past 3 months, how often did you have pain? Would you say never, some days, most days, or every day*?” Respondents who answered “Most days” or “Every day” were considered to have CP. High-impact CP was defined using the item: “*Over the past 3 months, how often did your pain limit your life or work activities? Would you say never, some days, most days, or every day?”* Respondents with CP who answered “Most days” or “Every day” were considered to have **High-Impact Chronic Pain (HICP)**; Respondents with CP who answered “never” or “some days” were considered to have **Lower-Impact Chronic Pain (LICP)**. Definitions of CP and HICP are aligned with the International Association for the Study of Pain (IASP) classification of chronic pain for International Classification of Diseases (ICD-11).^8^ Though CP and HICP constitute this study’s primary analytic focus, their logically nested relationship could be confusing. LICP is included in select reporting to facilitate contextualized interpretation of mean estimates, and to support representative, non-redundant visualizations of the U.S. adult population.

**PHQ-8 Score** was measured by summing the 8-item Patient Health Questionnaire (PHQ-8). The PHQ-8 asks respondents to answer: “*Over the last 2 weeks, how often have you been bothered by any of the following problems?”* **PHQ-1 Anhedonia:** “*Little interest or pleasure in doing things”* ; **PHQ-2 Sadness/Blues:** “*Feeling down, depressed, or hopeless”*; **PHQ-3 Sleep:** *“Trouble falling or staying asleep, or sleeping too much*”; **PHQ-4 Fatigue: “***Feeling tired or having little energy*”; **PHQ-5 Appetite:** “*Poor appetite or overeating*”; **PHQ-6 Self-Blame:** “*Feeling bad about yourself – or that you are a failure or have let yourself or your family down*”; **PHQ-7 Concentration: “***Trouble concentrating on things, such as reading the newspaper or watching television*”; and **PHQ-8 Psychomotor Function: “***Moving or speaking so slowly that other people could have noticed? Or the opposite – being so fidgety or restless that you have been moving around a lot more than usual*.” Responses are ordinal: (0) “Not at all,” (1) “Several Days,” (2) “More than Half the Days,” and (3) “Nearly Every Day.” **Positive** s**creens** were determined using the standard cut point for PHQ-8 (≥10).^22^ **Negative screens** included those with **absent/minimal symptoms** (PHQ scores 0-4) or **subclinical/mild symptoms** (PHQ-8 scores 5-9); **positive screens** include those with **moderate symptoms** (PHQ scores 10-14) or **severe symptoms** (PHQ-8 scores 15-24).^16^

Categorization of PHQ-8 items as somatic or cognitive/affective symptoms was aligned with literature:^23^ **somatic score** was calculated by summing items 3, 4, 5, and 8— sleep, fatigue, appetite, and psychomotor functioning; **cognitive/affective score** was calculated by summing items 1, 2, 6, and 7—anhedonia, sadness, self-blame, and concentration. For respondents with non-zero PHQ-8 scores, **somatic symptoms proportion** was calculated as the somatic symptom score divided by the PHQ-8 total score. Respondents with PHQ-8 scores of zero were coded as zero for the proportion.

### 2.3 Missing Data

A total of 1,009 observations (3.15% of data) were missing (responses of “Refused,” “Not Ascertained,” or “Don’t Know”) from PHQ-8, CP, or both. Considering study data characteristics of large sample size, small amount of missing data, and minimal expectation of precision and bias loss in the context of observational data, which suggest that deviations from the missing at random (MAR) assumption are unlikely to affect validity of estimates, complete case analysis was used.^24^ We evaluated means for key study variables comparing observations with missing data and those without missing data; consistent means provided additional support that the MAR assumption is justifiable.

### 2.4 Study design and analysis plan

Preliminary descriptive analyses examined prevalence of positive PHQ-8 screens by CP status, distributions of PHQ-8 scores and item-level responses grouped by CP and depression status **(Figure 1)** and assessed whether CP was associated with differences in the proportional contribution of somatic symptoms in PHQ-8 scores **(Figure 2)**.

**Figure 1.**
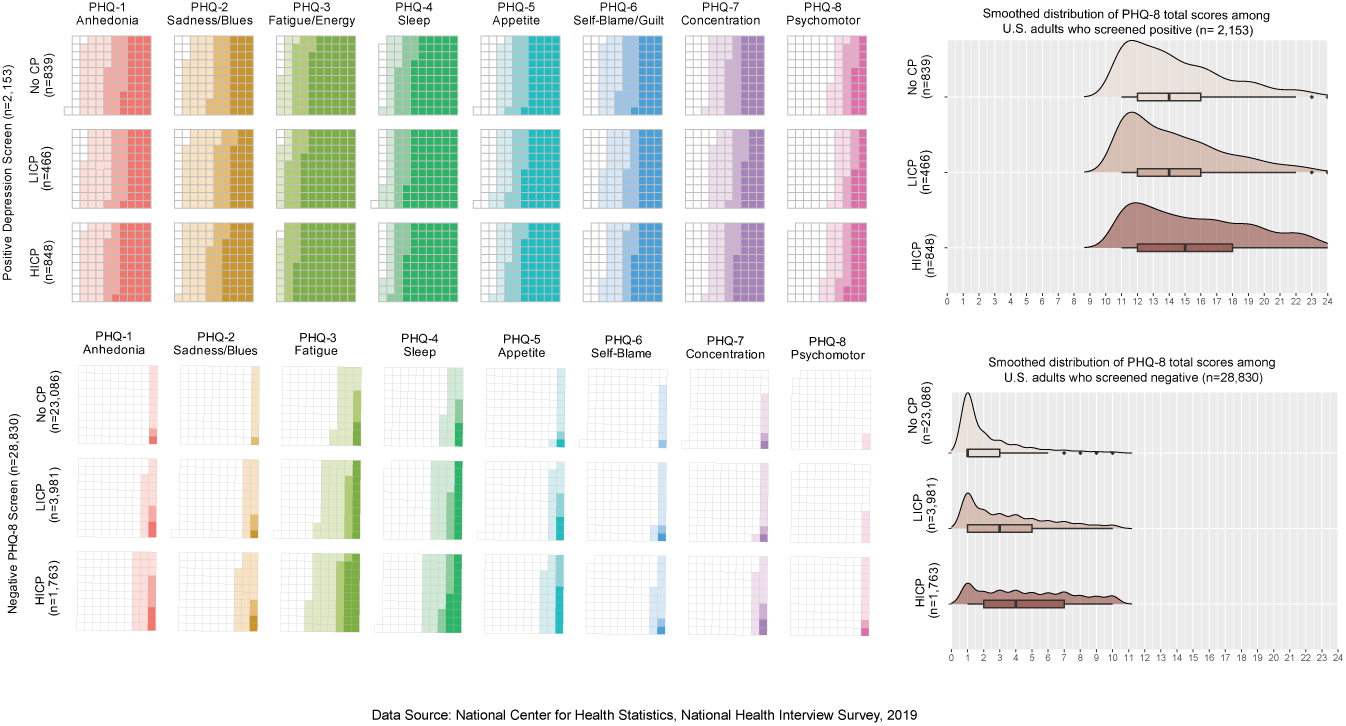
Item-level responses and total PHQ-8 scores of U.S adults, grouped by mutually exclusive and exhaustive combinations of PHQ-8 screening status and CP status. Abbreviations: NoCP= those without chronic pain; LICP = those with lower impact CP; those with HICP = high-impact chronic pain. Waffle plots(panels at left): each unit square in 10×10 grids represents 1% of subgroup. Color fill represents severity of each symptom; No color = “Not at all”, Lightest color = “Several days”, Medium Color = “More than half the days”, Darkest color = “Nearly every day”.

**Figure 2.**
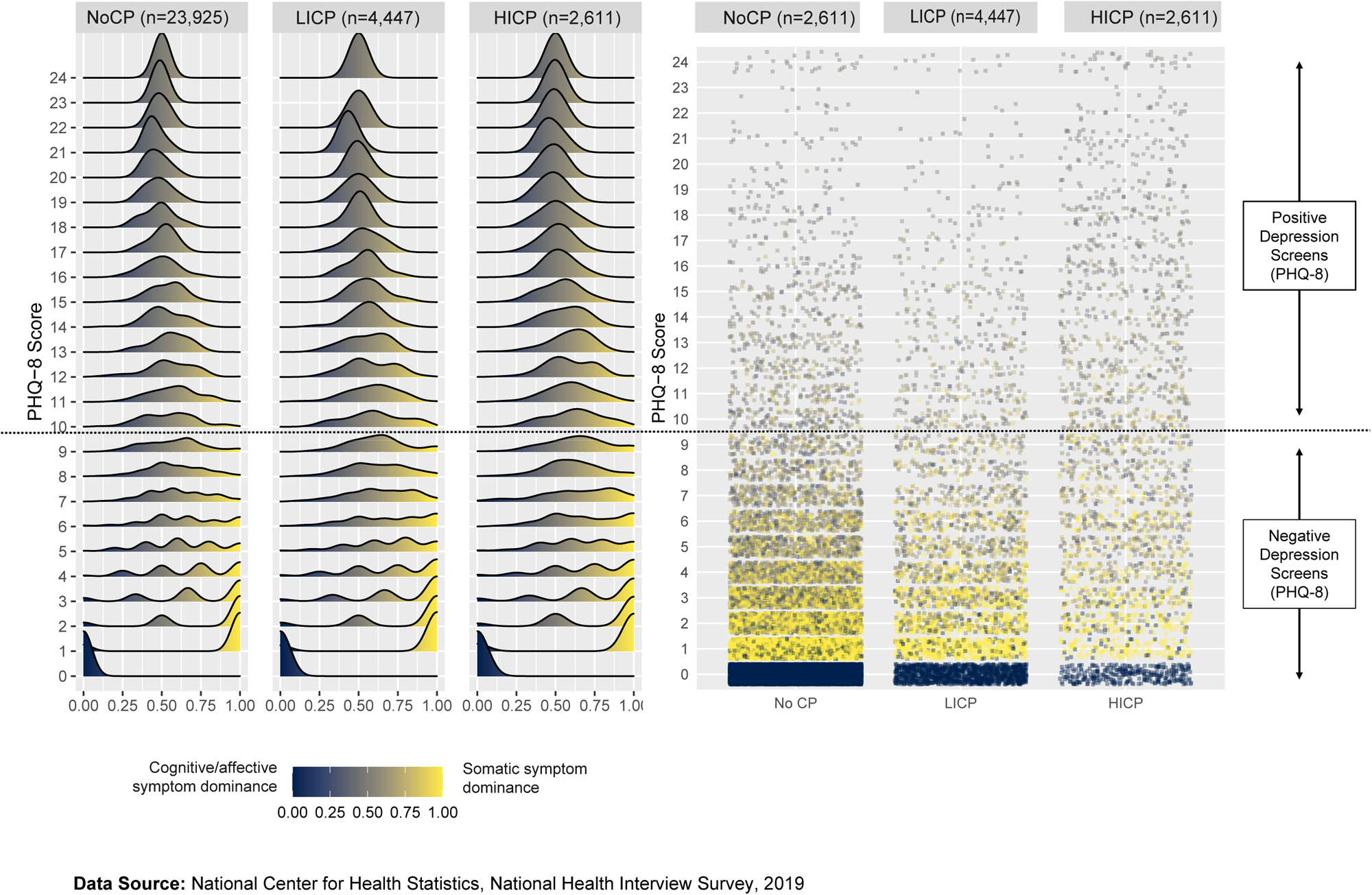
Smoothed density and scatter plots showing the proportion of somatic symptoms within PHQ-8 scores, grouped by mutually exclusive and exhaustive subgroups to facilitate representative visualization of the U.S. population.

Next, the reliability and cross-group equivalence and measurement invariance of PHQ-8 were evaluated, using the Chi-Square test statistic, Comparative Fit Index (CFI), Tucker-Lewis Index (TLI), Root Mean Square Error of Approximation (RMSEA), and Standardized Root Mean Square Residual (SRMR).

Reliability was assessed using Cronbach’s alphas and McDonald’s Omega coefficients; values greater than 0.80 were considered indicative of acceptable reliability **(Table 1)**.^25^ CFI and TLI values greater than 0.95 are indicative of good model fit; RMSEA and SRMR values lower than .05 indicate close fit and <.08 are indicative of reasonable model fit.^26,27^

**Table 1.**
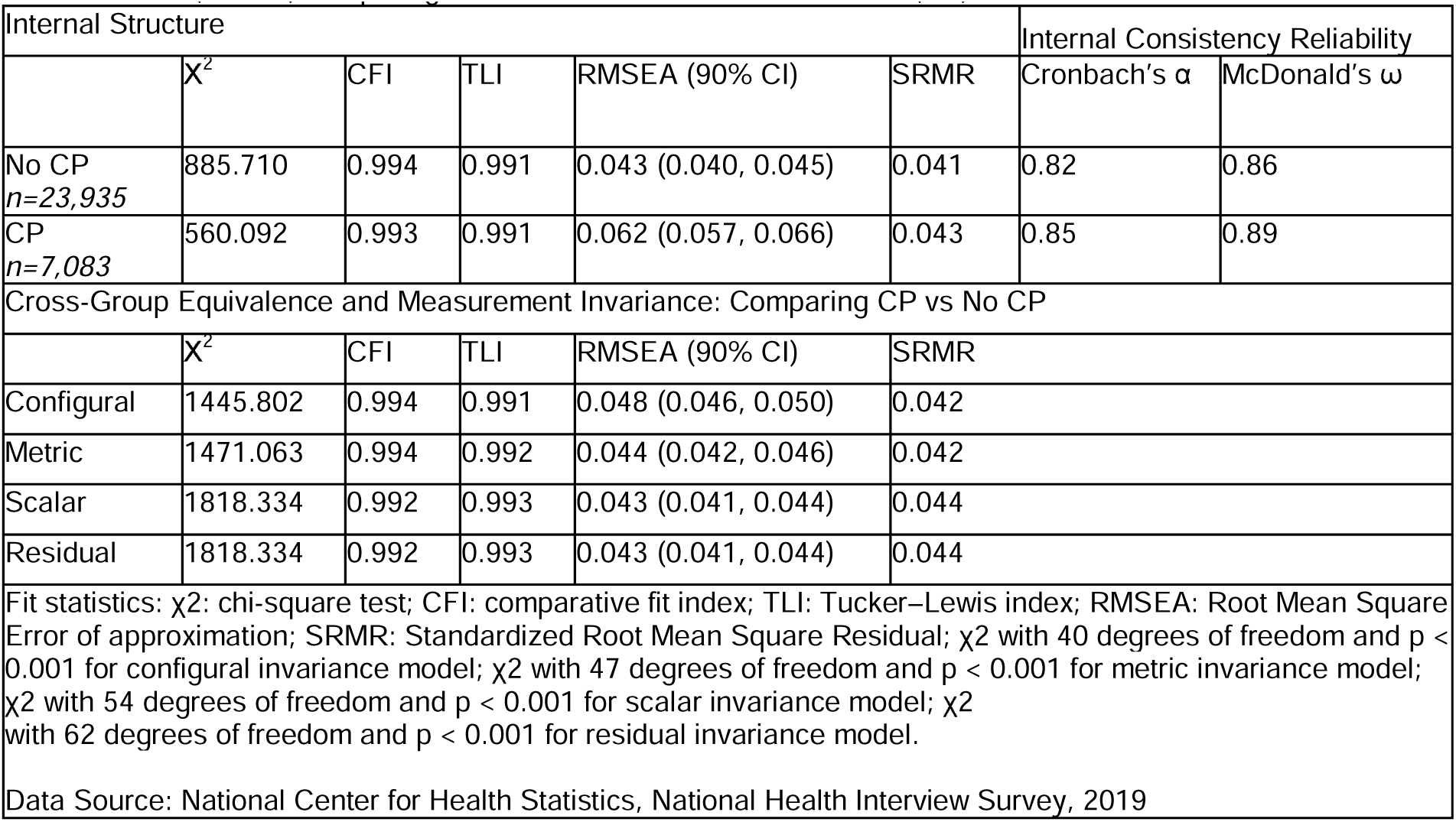
Internal structure, reliability, cross-group equivalence and measurement invariance of the Patient Health Questionnaire (PHQ-8) comparing those with and without Chronic Pain (CP).

| Internal Structure |  |  |  |  |  | Internal Consistency Reliability |  |
| --- | --- | --- | --- | --- | --- | --- | --- |
| | $\chi^2$ | CFI | TLI | RMSEA (90% CI) | SRMR | Cronbach's $\alpha$ | McDonald's $\omega$ |
| No CP<br>*n*=23,935 | 885.710 | 0.994 | 0.991 | 0.043 (0.040, 0.045) | 0.041 | 0.82 | 0.86 |
| CP<br>*n*=7,083 | 560.092 | 0.993 | 0.991 | 0.062 (0.057, 0.066) | 0.043 | 0.85 | 0.89 |
| Cross-Group Equivalence and Measurement Invariance: Comparing CP vs No CP |  |  |  |  |  |  |  |
| | $\chi^2$ | CFI | TLI | RMSEA (90% CI) | SRMR | | |
| Configural | 1445.802 | 0.994 | 0.991 | 0.048 (0.046, 0.050) | 0.042 |  |  |
| Metric | 1471.063 | 0.994 | 0.992 | 0.044 (0.042, 0.046) | 0.042 |  |  |
| Scalar | 1818.334 | 0.992 | 0.993 | 0.043 (0.041, 0.044) | 0.044 |  |  |
| Residual | 1818.334 | 0.992 | 0.993 | 0.043 (0.041, 0.044) | 0.044 |  |  |
| Fit statistics: $\chi^2$: chi-square test; CFI: comparative fit index; TLI: Tucker–Lewis index; RMSEA: Root Mean Square Error of approximation; SRMR: Standardized Root Mean Square Residual; $\chi^2$ with 40 degrees of freedom and $p < 0.001$ for configural invariance model; $\chi^2$ with 47 degrees of freedom and $p < 0.001$ for metric invariance model; $\chi^2$ with 54 degrees of freedom and $p < 0.001$ for scalar invariance model; $\chi^2$ with 62 degrees of freedom and $p < 0.001$ for residual invariance model. | | | | | | | |
| Data Source: National Center for Health Statistics, National Health Interview Survey, 2019 |  |  |  |  |  |  |  |

Cross-group equivalence and measurement invariance were assessed using a stepwise approach. First, configural invariance was evaluated to determine whether the factor structure was equivalent across participants with and without CP. Next, metric invariance was tested by constraining factor loadings to be equal across groups. Scalar (threshold) invariance was then examined by constraining both factor loadings and item thresholds to equality. Finally, residual invariance was assessed by additionally constraining residual variances to equality across groups **(Table 1).** The process was repeated for HICP and LICP **(Supplemental Table 1).**

Standardized factor loadings and item-discrimination parameters using graded response models were assessed (**Table 2).** Standardized factor loadings greater than 0.7 were indicative of “excellent” item performance; those greater than .5 and less than or equal to .7 were indicative of “good” item performance.^28^ Discrimination parameters greater than or equal to 1.70 were considered “quite satisfactory;” those greater than or equal to 1.35 and less than 1.70 were considered “good.”^29^ Item Response Theory (IRT) Test Information Functions (TIFs) and Item Information Functions (IIFs) were evaluated **(Figure 3)**. Analyses were completed using R version 4.4.2.; several packages were used: *ggplot*2, *survey, lavaan,* and *mirt*.

**Figure 3.**
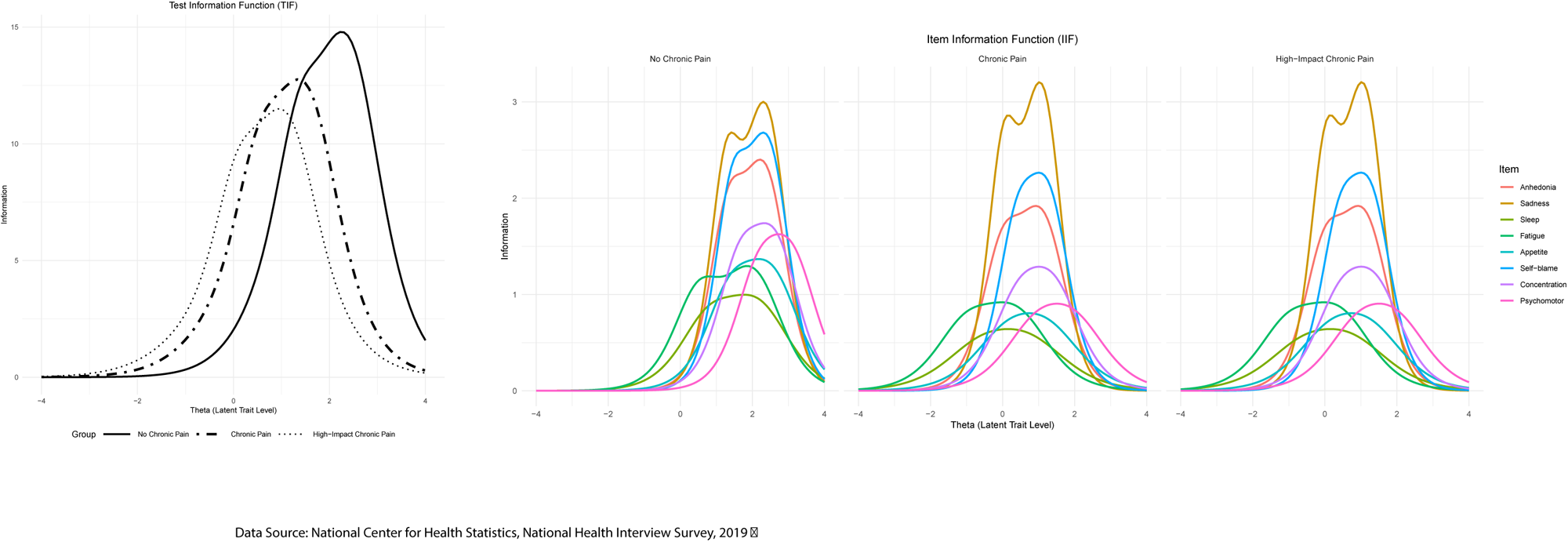
Patient Health Questionnaire (PHQ-8) Test information functions (TIFs) and item information functions (IIFs), comparing U.S. adults without chronic pain (CP), U.S. adults with CP, and U.S. adults with high-impact chronic pain.

**Table 2.** Confirmatory Factor Analysis (CFA) standardized item loadings and Item Response Theory (IRT) discrimination parameters in U.S. adults without chronic pain (NoCP), with chronic pain (CP), and with high-impact chronic pain (HICP).

| CFA Standardized Item Loadings |  |  |  |  |  |  |  |  |  |
| --- | --- | --- | --- | --- | --- | --- | --- | --- | --- |
|  |  | anhedonia | sadness | sleep | fatigue | appetite | self-blame | concentration | psychomotor |
| | n | $\lambda_1$ | $\lambda_2$ | $\lambda_3$ | $\lambda_4$ | $\lambda_5$ | $\lambda_6$ | $\lambda_7$ | $\lambda_8$ |
| NoCP | 23,925 | 0.841 | 0.875 | 0.714 | 0.755 | 0.744 | 0.839 | 0.783 | 0.759 |
| CP | 7,063 | 0.833 | 0.876 | 0.755 | 0.731 | 0.731 | 0.834 | 0.777 | 0.727 |
| HICP | 2,611 | 0.812 | 0.884 | 0.665 | 0.734 | 0.685 | 0.823 | 0.754 | 0.690 |

| IRT Discrimination Parameters |  |  |  |  |  |  |  |  |  |
| --- | --- | --- | --- | --- | --- | --- | --- | --- | --- |
|  |  | anhedonia | sadness | sleep | fatigue | appetite | self-blame | concentration | psychomotor |
| | n | $\alpha_1$ | $\alpha_2$ | $\alpha_3$ | $\alpha_4$ | $\alpha_5$ | $\alpha_6$ | $\alpha_7$ | $\alpha_8$ |
| No CP | 23,925 | 2.821 | 3.167 | 1.810 | 2.085 | 2.109 | 2.978 | 2.389 | 2.290 |
| CP | 7,063 | 2.710 | 3.144 | 1.528 | 1.931 | 1.883 | 2.829 | 2.244 | 1.951 |
| HICP | 2,611 | 2.523 | 3.277 | 1.442 | 1.737 | 1.615 | 2.724 | 2.044 | 1.712 |

**Data Source:** National Center for Health Statistics. National Health Interview Survey, 2019

## 3. RESULTS

### 3.1 Preliminary Descriptive Analyses

#### Comparing Prevalence of Positive PHQ-8 screens by CP status

The prevalence of positive PHQ-8 screens was 6.9% in the U.S. adult general population. U.S. adults with CP or HICP had significantly higher prevalence of positive screens compared to those without CP (NoCP): prevalence of positive screens was 3.5% in NoCP compared to 20.1% in CP— reflecting prevalence of 11.5% in LICP and 34.8% in HICP (all p-values <0.001). Compared to NoCP, positive screens were 5 times more prevalent in CP, and nearly 10 times more prevalent in HICP.

#### Comparison of PHQ-8 scores by CP status

*In the general population,* the mean PHQ-8 score was 2.48. Those with CP or HICP had significantly higher scores compared to those without CP. The mean score was 1.77 in NoCP compared to 5.25 in CP: reflecting means of 3.85 in LICP and 7.71 in HICP (all p-values <.001).

*Among those who screened negative,* the mean PHQ-8 score was 1.61; the mean score was 1.33 in NoCP compared to 2.91 in CP—reflecting means of 2.58 in those with LICP and 3.72 in those with HICP (all p-values <.001). Among those with negative screens, the distributions of PHQ-8 scores differed qualitatively across NoCP, CP, and HICP. Absent/minimal depressive symptoms (PHQ-8 scores 0-4) were the modal screening outcome across all groups, however absent/minimal symptoms were more prevalent in NoCP. The prevalence of absent/minimal symptoms was 90.88% in NoCP compared to 72.18% in CP — reflecting prevalence of 77.04% in LICP and 60.58% in HICP (all p-values <.001) **(Figure 1, lower right panel).**

*Among those who screened positive,* the mean PHQ-8 score was 14.29; the mean was 13.83 in NoCP compared to 14.61 in CP — reflecting means of 13.65 in LICP and 15.12 in HICP (all p-values <.001). Among those with positive screens, the distributions of PHQ-8 scores were qualitatively similar across NoCP, CP, and HICP **(Figure 1, upper right panel)**. While statistically significant CP-associated differences in mean PHQ-score were observed among those with positive screens, the magnitude of the difference was considerably attenuated in this group compared to those who screened negative. Moderate depressive symptoms (PHQ-8 scores 10-14) were the modal screening outcome across all groups, however severe symptoms (PHQ-8 scores 15-24) were disproportionately prevalent in CP. The prevalence of severe symptoms was 34.6% in NoCP and 43.7% in those with CP: reflecting prevalence of 33.9% in LICP and 49.3% in HICP (all p-values <.001).

#### Comparison of individual PHQ-8 items by CP status and screening status

*Among those who screened negative*, CP was associated with statistically and substantively significant increases in mean severity of the 8 individual items (scores ranging from 0 to 3): anhedonia (0.12 vs. 0.28), sadness (0.11 vs. 0.26), sleep (0.33 vs. 0.72), fatigue (0.41 vs. 0.89), appetite(0.14 vs 0.33) self-blame (0.09 vs. 0.18), concentration (0.08 vs. 0.18) and psychomotor functioning (0.03 vs. 0.08); the magnitude of the CP-associated differences was intensified in HICP (all p-values <.001).

However, *among those who screened positive,* CP-associated differences in symptom severity were statistically and substantively attenuated. CP was associated with statistically significantly (p<.001) increases in mean severity of sadness (1.65 vs. 1.78), sleep problems (2.24 vs. 2.37), and fatigue (2.33 vs. 2.60); mean differences in severity of anhedonia, appetite, self-blame, concentration, and psychomotor functioning did not reach statistical significance. Statistically significant increases in severity were observed in HICP for seven of the PHQ-8 items (p<.001); only concentration did not reach significance. We visualized response distributions for each item grouped by screening and CP status. (**Figure 1, panels at left**). It is worth noting that in all six combinations of screening status (negative vs. positive) and CP status (NoCP vs. LICP vs. HICP), the mean severity of sleep and fatigue was at least twice that of the mean severity of other items (p<.001 across all CP and screening status).

#### Proportional contribution of somatic symptoms

The proportion of somatic symptoms was calculated for all respondents, the distribution of the calculated proportions were visualized and compared across NoCP, LICP, and HICP **(Figure 2).** *Among those who screened negative,* dominance of either somatic symptoms or cognitive/affective symptoms was frequently observed, however, *among those who screened positive*, observations with balanced proportions of somatic and cognitive/affective symptoms are increasingly prevalent. This general pattern was uniform across NoCP, LICP, and HICP.

Specifically, *among those who screened positive,* the mean proportion of somatic symptoms was 0.55. The mean proportion was 0.53 in NoCP, compared to 0.55 in CP: reflecting proportions of .56 in LICP and 0.55 in HICP. The difference in somatic symptom proportions (0.019) was statistically significant (p < .008), given the large sample size, but the magnitude of the difference is trivial.

### 3.2 Reliability and Measurement Invariance

The PHQ-8 shows excellent internal consistency reliability; cross-group equivalence and measurement invariance are observed. The test of measurement invariance determines whether the CFA factor loading matrix is invariant across groups, i.e. whether the PHQ-8 items relate to the single latent trait of depression in the same way. Inspection of model fit indices showed that cross group equivalence and measurement invariance are supported in CP **(Table 1**), and HICP (**Supplemental Table 1**) at the configural (the overall factor structure), metric (factor loading), and scalar (item intercept) levels, suggesting that the PHQ-8 measures depression and functions in a similar way among U.S. adults without CP, with CP, and with HICP.

### 3.3 CFA Standard Factor Loadings and IRT Discrimination Parameters

CFA and IRT models are conceptually equivalent, yet the interpretation of standard factor loadings and discrimination parameters is complementary;^30^ therefore, we report both **(Table 2**). Higher standard factor loadings indicate a greater shared variance between the item and the underlying factor. All standardized factor loadings (λ) exceeded 0.660 across groups, indicating “very good” to “excellent” item performance with substantial shared variance between each PHQ-8 item and the underlying depression factor. The NoCP group demonstrated the strongest factor structure (λ: 0.714-0.875), with all eight items achieving excellent performance (λ ≥ 0.71). The CP group showed comparable results (λ: 0.727-0.876) with eight items reaching excellent performance levels, while the HICP group exhibited the most variability (λ: 0.665-0.884) with five items achieving excellent performance and three items achieving good performance. Sadness demonstrated the highest factor loading (λ: 0.875-0.884) consistently across groups; sadness was the strongest indicator of depression regardless of CP status.

IRT discrimination parameters reflect the ability of each item to discriminate among people with differing *severity* of depression. All items demonstrated “good” to “quite satisfactory” discrimination across groups, with parameters (α) ranging from 1.442 to 3.277. Most items achieved quite satisfactory performance (α ≥ 1.70), across groups, indicating highly effective differentiation of severity between individuals. Sleep achieved relatively lower, though still “good” (α ≥ 1.35) discrimination ability in CP and HICP, while appetite, fatigue, and psychomotor functioning achieved "good" discrimination in HICP. This suggests that these somatic symptoms may be relatively less discriminant to identify different levels of depression among those with HICP; it is useful to note that this does not suggest problems with comparing groups on the same depression trait dimension.^30^

Evaluation of TIFs show that the PHQ-8 had acceptable measurement precision across different severity levels; IIFs are consistent across groups, and sadness (PHQ-2) again demonstrated the highest ability to detect differing levels of severity **(Figure 3).**

## 4. DISCUSSION

This representative population study (n= 30,983) demonstrated that the PHQ-8 reliably and equivalently measures depressive symptoms in U.S. adults with and without CP. Study results should enhance confidence among clinicians and researchers that the PHQ-8 is suitable to screen for depressive symptoms in the context of CP and that the interpretation of screening results is straightforward. Further, these results suggest that comparison of PHQ-8 total scores or individual PHQ-8 items between those with and without CP is analytically appropriate. Finally, findings persuasively suggest that the very high prevalence of positive PHQ-8 screens commonly reported among those with CP reflect true differences in the burden of clinically significant depressive symptoms and are not measurement artifacts.

Descriptive analyses, fit statistics, and results from tests of measurement invariance and cross group equivalence were consistent across groups. Factor loadings and item discrimination coefficients were likewise uniform and no differential item functioning was observed. Sadness had the highest factor loadings and was the most discriminant across groups, corroborating its importance in differentiating depressive symptoms and their severity regardless of CP status.

As expected, CP was associated with startling disparities in the prevalence of positive PHQ-8 screens for clinically significant depressive symptoms. Descriptive analyses of representative data and comparison between focal and non-focal groups did not suggest any need for qualification or nuance related to this finding. The prevalence of positive PHQ-8 screens was 3.5% in those without CP, 20.1% in those with CP, and 34.8% in those with high-impact chronic pain (HICP), representing 5-to 10-fold increases in the prevalence of unmet mental health needs. This study did not find evidence to suggest somatic (nor any other) symptoms may exert undue influence on the depression screening outcomes of those with CP compared to others. Consistent with prior studies, those with CP were observed to have elevated severity of several somatic items including fatigue and sleep disturbance when compared to the severity of other PHQ-8 items. However, comparison of focal and non-focal groups revealed that this finding is not unique to those with CP and that the same cross-item pattern is also readily observed among those without CP.

We then evaluated whether CP-associated differences existed in the proportional contribution of somatic symptoms, holding PHQ-scores constant. Cross-group differences between those with and without CP were not observed. Proportional dominance of somatic symptoms was frequently observed in those who screened negative on PHQ-8, while balanced contributions of somatic and cognitive/affective symptoms were typical among those who screened positive. The mean proportion of somatic symptoms decreased linearly as PHQ-8 score increased.

CP was associated with statistically significant increases in severity of all individual items and of total PHQ-8 scores. However, we suggest cautious interpretation of this difference as CP-associated differences in severity were disproportionately driven by those who screened negative on PHQ-8. Among those who screened positive, cross-group differences in severity of individual items and of PHQ-8 scores were attenuated or eliminated, however, significant if substantively attenuated differences were preserved in the context of HICP for the majority of items.

This study has limitations. First, we acknowledge that some individuals are not reached by the NHIS, including U.S. persons with no fixed address, persons living on military bases, and persons living in long-term care facilities or correctional facilities. Second, though the PHQ-8 items administered by NHIS are identical to those administered in clinical practice, it is still possible that screening data collected by NHIS could be less accurate than screening data collected in the context of a clinical encounter.

For several decades, the binary opposition of pain and depression has been misaligned with modern scientific understanding about the biopsychosocial nature of pain; scientific consensus is coalescing that dualistic assignment of symptoms to *either* CP *or* depression could be counterproductive.^3,14^ The literature is clear that addressing the unmet mental health needs of those with CP is associated with better clinical prognoses, global functioning, and quality of life. Yet, emerging research demonstrates that those with CP face end-to-end disparities across the mental health patient journey—including increased prevalence of mental health needs, reduced likelihood that depressive symptoms or other mental health needs will be identified, and reduced likelihood of receiving needed mental health support.^4^ Those who screen positive on PHQ-8 benefit from equitable access to information, resources, and mental health support, regardless of chronic pain status. Person-centered clinical communication, broad implementation of accessible CP-informed mental health care, and interventions to directly target CP-associated mental health stigmas are needed.

## Data Availability

No original data is produced by this study, the publicly available data set is available on the National Health Interview Survey websites
https://www.cdc.gov/nchs/nhis/documentation/2019-nhis.html

## Contributors

All authors had full access to all study data and take responsibility for the integrity of the data and the accuracy of the data analysis. JDLR, GC, and RA conceived and designed the study. JDLR, GC, and KH directly accessed, verified, and analyzed the study data reported in the manuscript. All authors, JDLR, GC, KH, CM, and RA interpreted the data, drafted the manuscript, and critically revised the manuscript for important intellectual content. JDLR supervised the project.

## Data sharing statement

All data from the National Health Interview Survey are publicly available on the website of the National Center for Health Statistics. Code is available at DOI: 10.5281/zenodo.16954664.

## Acknowledgements

This study was supported in part by the Comprehensive Center for Pain & Addiction, University of Arizona, Tucson, AZ, USA. RVA’s time is funded by NIH K23HD104934. Funders had no role in the design and conduct of the study; collection, management, analysis, and interpretation of the data; preparation, review, or approval of the manuscript; nor in the decision to submit the manuscript for publication.

