## Supplemental Table 1 for "Reliability and equivalence of the 8-item version of the Patient Health Questionnaire (PHQ-8) to screen for depressive symptoms in adults with chronic pain: A representative U.S. population study"

**Supplemental Table 1.** Internal structure, reliability, cross-group equivalence and measurement invariance of PHQ-8 comparing those with and without high-impact chronic pain (HICP).

|  | | **Χ^2^** | **CFI** | **TLI** | | | **RMSEA (90% CI)** | **SRMR** | **Cronbach’s α** | **McDonald’s ω** |
| --- | --- | --- | --- | --- | --- | --- | --- | --- | --- | --- |
| **No HICP**  *n*=28,407 | | 300.019 | 0.991 | 0.987 | | | 0.056 (0.051, 0.071) | 0.049 | 0.81 | 0.86 |
| **HICP**  *n=2,611* | | 309.754 | 0.991 | 0.987 | | | 0.075 (0.067, 0.082) | 0.050 | 0.85 | 0.89 |
| **Cross-Group Equivalence and Measurement Invariance: Comparing HICP vs No HICP** | | | | | | | | | | |
|  | **Χ^2^** | | **CFI** | | **TLI** | **RMSEA (90% CI)** | | | **SRMR** | |
| **Configural** | 609.774 | | 0.991 | | 0.987 | 0.064 (0.059, 0.068) | | | 0.049 | |
| **Metric** | 628.171 | | 0.991 | | 0.989 | 0.059 (0.055, 0.063) | | | 0.050 | |
| **Scalar** | 654.731 | | 0.990 | | 0.991 | 0.052 (0.048, 0.056) | | | 0.050 | |
| **Residual** | 654.731 | | 0.990 | | 0.991 | 0.052 (0.048, 0.056) | | | 0.050 | |
| Fit statistics: χ2: chi-square test; CFI: comparative fit index; TLI: Tucker–Lewis index; RMSEA: Root Mean Square Error of approximation; SRMR: Standardized Root Mean Square Residual; χ2 with 40 degrees of freedom and p < 0.001 for configural invariance model; χ2 with 47 degrees of freedom and p < 0.001 for metric invariance model; χ2 with 54 degrees of freedom and p < 0.001 for scalar invariance model; χ2 with 62 degrees of freedom and p < 0.001 for residual invariance model.  **Data Source:** National Center for Health Statistics, National Health Interview Survey, 2019 | | | | | | | | | | |
